# The dark side of streaking: Examining the backfire potential of run streaking in recreational runners who broke a long-term streak

**DOI:** 10.1101/2024.12.26.24319676

**Authors:** Erin E. Ingalls, Nicholas Larade, Gozde Ozakinci, Gabriela Tymowski-Gionet, Lena Fleig, Stephan U. Dombrowski

**Affiliations:** Faculty of Kinesiology, University of New Brunswick, Canada; Division of Psychology, University of Stirling, Scotland; Department of Psychology, Medical School Berlin, Germany

**Keywords:** Streaking, behaviour change technique, backfire potential, running, physical activity

## Abstract

**Background:** Run streaking is running on consecutive days for a minimum of one mile per day. Despite its benefits for supporting habit formation and long-term behaviour change, some streak runners report potential unintended negative consequences of run streaking. The aim of the study is to examine the backfire potential of run streaking in recreational runners who ended a long-term streak.

**Methods:** Qualitative semi-structured interviews with 17 recreational adult runners (10 male, 6 female, 1 other gender). All runners ended a run streak of ≥100 consecutive days. Transcripts were analyzed using a hybrid deductive–inductive thematic analysis.

**Results:** Prior to streak cessation, some runners felt streak-related inconveniences and ran with injury to prolong the streak. Immediate consequences following the end of a streak included feelings of sadness, anger, disappointment and relief. Several run streakers described a ‘grieving process’ in the weeks and months following streak cessation. Unintended negative consequences were amplified in runners with higher levels of streak attachment. All physically capable runners continued to run regularly with most starting a new streak and all voiced positive views towards run streaking despite their streak ending.

**Conclusion:** Run streaking as a behaviour change technique has small backfire potential in some runners. Ending a long-term run streak can lead to short-term negative affect which can develop into experiences of grief, particularly in those with high levels of streak attachment. No long-term negative consequences were reported. All participants perceived run streaking as positive overall and remained physically active following the end of their long-term streak.

## INTRODUCTION

Regular engagement in physical activity leads to mental and physical benefits and supports the primary and secondary prevention of various chronic diseases such as diabetes mellitus, cancer, obesity, hypertension, and depression (1, 2). The Canadian 24-Hour Movement Guidelines suggest that adults engage in 150 minutes of aerobic moderate-to-vigorous intensity physical activity each week (3). However, despite the well-documented benefits of regular physical activity, many people do not meet these guidelines. In Canada, for example, 51% of Canadians have been estimated to not accumulate 150 minutes of physical activity per week (4). A potential aerobic physical activity behaviour that can help individuals reach physical activity guidelines is running.

Running is the most fundamental vigorous-level activity and benefits overall physical and mental health (5). A systematic review of 14 studies from six prospective cohorts with a pooled sample of 232,149 participants found that running participation was associated with 27%, 30%, and 23% lower risk of all-cause, cardiovascular, and cancer mortality, respectively, compared to no running (6). A large prospective cohort study on running and longevity examined 55,137 adults over a 15 year average follow-up period estimated that runners (approximately 24% of the cohort) lived an average of three years longer than non-runners (7). Moreover, a longitudinal study of 244 recreational runners that examined the relationship between running and psychological wellbeing (8) found that runners thought better about themselves and their lives, and the better they felt affectively, the more frequently they ran and the more distance they ran during a week. Frequency of running seemed a more reliable predictor of well-being than the distance covered, suggesting that regular running episodes might benefit psychological wellbeing in runners.

Sustained physical activity performance is required to obtain and maintain the health benefits associated with being active. However, running cessation is common in recreational runners (9). According to a prospective cohort study including 774 participants, lack of motivation and injury are common variables associated with the discontinuation of running (10). Conversely, self-regulation skills and intrinsic motives were reported as key antecedents of running behaviour in a systematic review including 58 studies that examine psychological correlates of recreational running (11). A longitudinal qualitative study of 20 new runners identified that having a meaningful reason to run (such as those related to identity, values, special memories, relationships, enjoyment or a personal goal) and experiencing progress as important for maintaining running behaviour for 6 to 12 months (12).

The development of habits has been theorized to play a significant role in maintaining behaviour change (13). Repetition is a central component in the formation of habits (14). It has been estimated that habit formation takes an average of 66 days, with a range of 18-254 days (15) and repetition has been theorised as a key component to support the formation of habits particular in the early days of habit formation (16, 17). Engaging in a behaviour frequently—such as every day—facilitates repetition and habit formation.

Behavioural streaking is the performance of a pre-specified behaviour consecutively in regular intervals—usually daily—without taking a break, while tracking the number of repeated performances. Streaking as a behavior change technique has been examined in the context of marketing (18–20), but research on behavioural streaking within the health context is rare. Despite its frequent use in the general population and within marketing contexts, behavioural streaking as a technique is absent from the behaviour change technique (BCT) taxonomies, lists, and compendiums (21–24).

The application of behavioural streaking within the recreational running is typically defined as running on consecutive days for a minimum period of time or distance: typically at least one mile (1.61 kilometres), at any pace, at any place, and monitoring the number of days (25). Several recreational runners have adopted this technique and report successfully maintaining their daily running behaviour. Recent research examined run streaking as a BCT in recreational runners (26). The qualitative study including 21 recreational runners reported that run streaking may lead to habit formation of running behaviour. Run streaking was found to engage a variety of additional psychological mechanisms which support sustained behavioural performance over time, such as motivation, self-regulation, and identity. However, some runners reported unintended consequences of using this technique such as injury or stress.

Behaviour change interventions have the potential to lead to unintended negative consequences, sometimes referred to as a backfire (27). Potential unintended negative consequences of BCTs need to be understood to ensure an awareness of their inherent ‘backfire potential’ when using a technique. Backfire potential may be conceptualised as the likelihood of technique engagement leading to unintended negative consequences across different behaviours, populations, and contexts. Frameworks conceptualising how interventions targeting behaviour and behaviour-related change may lead to unintended negative consequences have been proposed (28, 29). The compendium of self-enactable techniques lists 123 techniques, with seven techniques accompanied by expert generated suggestions on **‘**possible adverse effects’ (21). For example, the technique ‘behavioral goal setting’ was accompanied by the mentioning of the possible adverse effect that failure to achieve goals may reduce feelings of competence and lead to disappointment. A better understanding of potential unintended negative consequences of techniques might help successful technique implementation and avoid BCT backfiring.

Based on previous streaking research (26) it can be hypothesised that streaking in the context of running might backfire in three areas leading to potential unintended negative consequences. First, run streaking might lead to health compromising effects such as increasing of psychological stress, or worsening of injury or lack of recovery from physical or mental challenges which may or may not be related to running. Second, the cessation of a run streak might negatively impact psychological wellbeing particularly for those run streakers who accumulated a run streak over several months and/or years. Third, run streak cessation might lead to an abandoning of running behaviour specifically or physical activity in general if the motive of behavioural engagement is solely motivated by upholding the streak.

The overall aim of this study is to examine the backfire potential of using run streaking as a BCT with a focus on recreational runners who ended a long-term run streak, as this population is most likely to have experienced potential unintended negative consequences of run streaking. The specific objectives were to i) explore perceptions during the lead up period prior to streak cessation, ii) reflect on the immediate consequences of run streak cessation, and iii) examine longer-term consequences of run streak cessation.

## METHODS

### Study Design

This study used a qualitative design. Data were collected between 13. November and 19 December 2023 through 17 semi-structured interviews with recreational streak runners who identified as male (n=10), female (n=6) and another gender (not disclosed to preserve anonymity, n=1) who ended a run streak of a minimum of 100 days. Interviews were conducted online, averaging 28.4 minutes in length (range = 16-52 min). Interviews were conducted by a researcher (EI) and supervised by a health psychologist (SD). Following verbal consent, each interview was recorded, transcribed, and anonymized prior to analysis. This study received approval from the University of New Brunswick Research Ethics Board and is on file as REB 2023-12B.

### Inclusion Exclusion Criteria

Participants were eligible for the study based on three criteria. Individuals were recruited who 1) were 19 years of age or older, 2) had completed a minimum of a 100-day streak (defined as running a minimum of one mile every day), which ended by choice or circumstance, and 3) were a recreational runner (i.e. not being paid for their running activities). There were no geographical restrictions as interviews were conducted virtually. Interviews were completed in English; thus, the ability to communicate in English was required for participation.

### Recruitment

Participants were recruited using an online flyer posted to a streak running Facebook group. The poster included a large title reading “Have you ever broken your run streak?” as well as the list of inclusion and exclusion criteria. Individuals interested in volunteering were encouraged to reach out to a member of the research team via Facebook or the email provided.

### Procedures

This project used a qualitative approach by interviewing recreational runners who had previously used the technique of run streaking. Interviews were conducted using Microsoft Teams or Zoom (participant preference). Prior to the interview, participants received an email containing information on the purpose and methods of the study. During the interview, participants were re-read the information contained in the email. Verbal consent was obtained from the individual for their participation in the study. Pre-selected interview questions remained consistent between each participant, and demographics were taken in the end (see Appendix A for the topic guide).

### Analysis

Interview transcriptions were anonymized and underwent abductive thematic analysis (30), moving between inductive and deductive coding (31) using Atlas.ti. This allowed for the construction of unanticipated themes and knowledges, as well as exploration through established temporal concepts (i.e. lead-up period prior to the streak ending; immediate consequences of the run streak ending; and longer-term consequences following the ending of the run streak).

Transcripts were first coded by one author using a deductive approach in line with the pre-specified research objectives based on broad general temporal sequence of the phenomenon under investigation and its inherent psychological processes.

Within the broad inductive temporal focus, codes were generated inductively to identify additional themes. Coding included identification of additional themes, which were discussed by the research team, defined, and refined. All codes were checked by a health psychologist (SD) to ensure that the transcriptions were consistently coded, as part of regular meetings during the analysis phase and discussed among the research team.

All interviews were conducted by a research student (EI) who did not engage in streak running at the time of interviews. This allowed participants to elaborate on their run streaking views and experiences in detail without assuming prior knowledge of the interviewer. The research team consisted of individuals who identified as runners without engaging in streak running (GO, GTG) streak runners (NL, SD), and those who identified as a physical activity streak in a different domain (i.e. yoga)/ (LF).

## RESULTS

### Participants

Seventeen runners (10 male, 6 female, 1 other gender) participated in this study (see Table 1). Participants were aged 20-29 (n=1), 30-39 (n=2), 40-49 (n=6), 50-59 (n=2), 60-69 (n=4), and 70+ (n=2). Based on self-reported height and weight, participants were placed into BMI categories: 18.5-24.9 (n=9), 25-29.9 (n=7), and 30-39.9 (n=1). Participants reported their marital status as being married (n=11), in a partnership (n=3), or single (n=3) and most came from the USA (n=11), followed by Canada (n=5), and Scotland (n=1). Ethnicities were reported as Caucasian (n=16) and Chinese (n=1) and participants were employed (n=12), unemployed (n=1), or retired (n=4).

**Table 1:** Participant demographics.

| **ID** | **Age Range (years)** | **Running experience (years)** | **Broken streak length (days)** | **Gender** | **BMI Category** | **Marital Status** | **Country** | **Ethnicity** | **Employment Status** |
| --- | --- | --- | --- | --- | --- | --- | --- | --- | --- |
| 1 | 60-69 | 21-30 | 501-1000 | M | 25.0-29.9 | Married | USA | White | Retired |
| 2 | 50-59 | 11-15 | 1001-2000 | M | 25.0-29.9 | Married | USA | White | Employed |
| 3 | 30-39 | 5-10 | 501-1000 | F | 18.5-24.9 | Partnership | Canada | Chinese | Employed |
| 4 | 60-69 | 21-30 | 501-1000 | M | 25.0-29.9 | Single | USA | White | Employed |
| 5 | 60-69 | 21-30 | 1001-2000 | F | 18.5-24.9 | Partnership | Scotland | White | Employed |
| 6 | 40-49 | 16-20 | 366-500 | O | 18.5-24.9 | Single | Canada | White | Employed |
| 7 | 40-49 | 11-15 | 100-365 | M | 25.0-29.9 | Partnership | Canada | White | Employed |
| 8 | 30-39 | 5-10 | 100-365 | M | 18.5-24.9 | Single | Canada | White | Employed |
| 9 | 40-49 | 16-20 | 1001-2000 | F | 18.5-24.9 | Married | USA | White | Employed |
| 10 | 60-69 | 40+ | 5000+ | M | 18.5-24.9 | Married | USA | White | Retired |
| 11 | 40-49 | 16-20 | 3001-5000 | M | 25.0-29.9 | Married | USA | White | Employed |
| 12 | 50-59 | 40+ | 1001-2000 | M | 18.5-24.9 | Married | Canada | White | Employed |
| 13 | 20-29 | 5-10 | 501-1000 | F | 25.0-29.9 | Married | USA | White | Employed |
| 14 | 40-49 | 16-20 | No data | F | 18.5-24.9 | Married | USA | White | Unemployed |
| 15 | 70-79 | 40+ | 2001-3000 | M | 18.5-24.9 | Married | USA | White | Retired |
| 16 | 70-79 | 40+ | 366-500 | M | 18.5-24.9 | Married | USA | White | Retired |
| 17 | 40-49 | 11-15 | 1001-2000 | F | 30.0+ | Married | USA | White | Employed |

Note: BMI= Body Mass Index (kg/m<sup>2</sup>), M= Male, F= Female, O = other gender (not disclosed to preserve anonymity).

Runners reported a range of general running experiences, including between 5-10 years (n=3), 11-15 years (n=3), 16-20 years (n=4), 21-30 years (n=3), and 40+ years (n=4). The length ranges of the broken streak were 100-365 days (n=2), 366-500 days (n=2), 501-1000 days (n=4), 1001-2000 days (n=5), 2001-3000 days (n=1), 3001-5000 days (n=1), 5000+ days (n=1) and unknown (n=1).

### Introduction of Themes and sub-themes

Figure 1 outlines the themes and sub-themes constructed in the process of data analysis. The study examined run streakers who had been engaging in a long-term run streak whose streak ‘broke’. A key underpinning theme that emerged when examining the experiences of long-term streaks ending which permeated subsequent themes and sub-themes identified was the notion of the streak becoming ‘a thing’ in many runners. Rather than being perceived as a change strategy one uses to support running behaviour, or as a grammatical noun adjunct (‘run streak’), runners behaved as if the streak itself was a separate external ‘thing’, which in some cases exhibited properties of living organisms. This ‘biomorphism of streaking’ was reflected in the language used around streaking, including expressions such as ‘*keeping the streak alive’*, ‘*streak saver*’. ‘*a streak breaking/ snapping/ dying*’ or ‘*losing a streak*’. The runner and their streak, over time, develop a relationship, whereby the continued investment in the streak by the runner helps the streak to continue existing and growing in size or age, and the streak in turn helps the runner fulfil their own unique needs such as engaging in physical activity or attaining a sense of daily achievement. The sub-themes in relation to the three temporal phases of long-term streak cessation (i.e., lead up period, immediate consequences, and longer-term consequences) were then developed based on the notion of the streak as a (living) thing.

**Figure 1.**
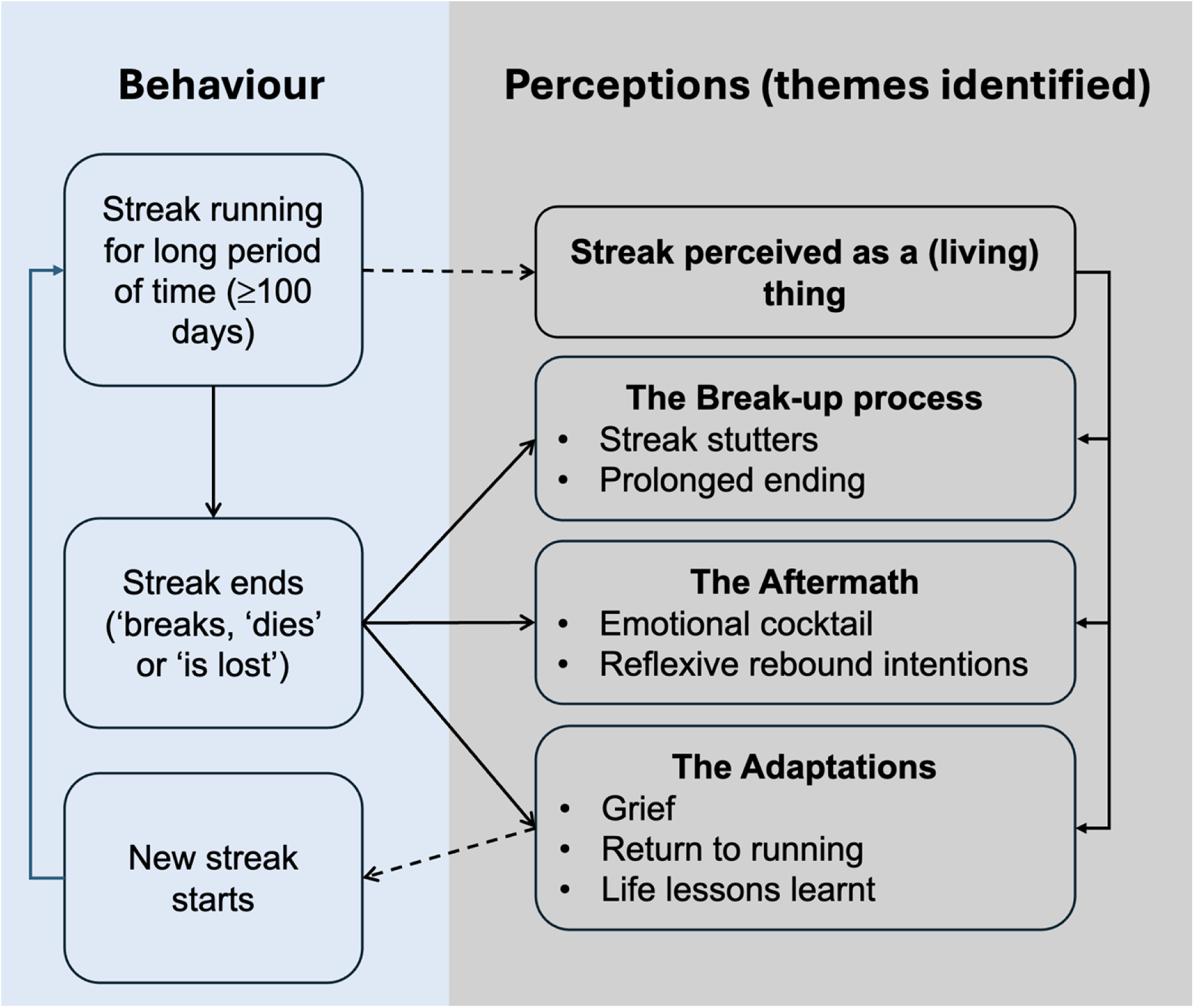
Framework depicting themes and corresponding sub-themes associated with long-term streak cessation.

The lead-up period related sub-themes were developed under the heading of ‘*the break-up process*’ and included two sub-themes: ‘*streak stutters*’ and ‘*prolonged ending’*. The immediate consequences related sub-themes were developed under the heading of ‘*the aftermath*’ and included two sub-themes: ‘*emotional cocktail*’ and ‘*reflexive rebound intentions*’. The longer-term consequences were developed under the heading of ‘*the adaptations*’ and included three sub-themes: ‘*grief*’, ‘*return to running’*, and ‘*life lessons learnt*’.

### The Break-up Process

In the lead up period to streak cessation, most runners outlined undergoing a process where the attitude towards their streak changed which ultimately led to the ending of their streak. The two sub-themes identified were: ‘*streak stutters*’ and ‘*prolonging of ending’*.

#### Streak stutters

‘*Streak stutters*’ refer to the repeated experience of inconveniences in the maintenance of run streaking. The run streak had the potential to induce physical strain, social conflict, and cognitive burden in the streak runner. The degree to which a runner perceived their streak to be an inconvenience was variable. Some reported minor stress associated with their streak, which were seen as manageable and did not interfere with streak maintenance.

> *“I wouldn’t say it was a burden. There is usually one single day, a single event [that feels like a burden], and then for the next three months, no burden at all, and then another single event that I would have to work around.”* Participant 15, Male.

Some described periods of inconvenience and burden when maintaining their run streak. When runners’ goals changed or were no longer being met, streak maintenance shifted from an overall enjoyable experience to one filled with burden.

> *“If [the streak] got to feel like a daily chore, then it defeated the purpose of trying to be fitter. It devolved into just sort of going out and doing the minimum so that I could maintain the streak. It was no longer achieving the objective that I’d set out for myself.”* Participant 12, Male.

During periods of streak stuttering where run streaking was not perceived as seamless and enjoyable, runners often reported reflecting on the purpose of their run streaking. These reflections and adverse experiences of run streaking took part whilst still practicing the daily running.

> *“Mentally, you can be extremely tired, worn out, and it can be wearing on you. Why am I doing this? […] Why do I keep doing this?”* Participant 1, Male.

#### Prolonged Ending

‘*Prolonged ending*’ refers to runners engaging in streak extension measures to avoid or delay streak cessation for as long as possible. The period of prolonged ending typically started following the experience of streak stutters which became more frequent or more intense over time, typically though injury or medical conditions unrelated to running. Runners frequently reported a continuation of running despite pain and were inclined to ‘*run through injury*’ to trying and maintain their streak.

> *“I certainly ran through some things that I would have normally taken a break for just because I wanted to keep my streak going”.* Participant 6, Other Gender.

Multiple runners reported injuries attained through running that were not given time to heal because of one’s persistence in the streak.

> *“Sometimes you can keep running when you have a bad injury and you say, ‘well, I’ll just run through it, I’ll just gut it out’, and you can do damage that makes it harder to heal from that injury or prolong the recovery of that injury.”* Participant 1, Male.

In addition to acute injuries attained through running, runners highlighted the long-term ‘wear and tear’ that their body experienced and age-related physical decline. Despite pain, runners remained dedicated to the continuation of their streak and there was a degree of normalization of injury for some runners experiencing persistent pain.

> *“I literally hurt all the time, every day something different hurts a little bit and I feel like that’s going to be unavoidable [when streaking].”* Participant 8, Male.

The prolonged ending period was often characterised by streak care behaviours (i.e., use of problem-oriented coping strategies) and goal-adjustment strategies such as reducing running intensity, time, or distance not leading to the desired outcomes anymore. When experiencing persistent periods of pain whilst continuing to engage in streak running, runners often reported engaging in a ‘streak saver’; a one-mile run, often completed at a slower than normal pace, solely done with the intention of ‘keeping the streak alive’. Rather than ending a streak when experiencing pain (i.e. avoidance behaviour), runners practiced pain endurance and engaged in ‘streak savers’ (i.e. running the minimum distance of one mile), to prolong the end of the streak.

> *“And I’m almost in tears. I’m like I can’t do this anymore, I just can’t. Like I can not do this physically anymore, it was like a month of painful one-milers.”* Participant 11, Male.

When surgery was required for running-related injuries, the experienced pain could become strong enough to force cessation of the streak before a scheduled surgery.

> *“At that point there was no way I was going to keep doing that to myself every day. It was pure agony basically, to go out and run.”* Participant 10, Male.

Some runners required surgery for non-running related reasons, and often reported the continuation of their streak up until the day of their scheduled surgery.

> *“I ran everyday up until the day of my surgery… so I stayed up until midnight the night before my surgery, ran, and then went to bed, so I could get that one last day in before my surgery.”* Participant 17, Female.

The experience of a final run before a scheduled ending, was viewed by some runners as a moment to celebrate the accomplishment of their streak.

> *“Well in my case it was kind of a celebratory run, that last run, because I knew it was the end of my streak”.* Participant 15, Male.

Others, however, felt more sadness in the run up to a scheduled streak ending and when executing what was known to be their final days of their streak.

> *“So leading up to that, I actually got really sad and I kind of didn’t give my last runs my all. I was just, I mean, I did, but I didn’t. I just knew it was the end.”* Participant 9, Female.

Runners with a strong attachment to their streak often delayed the ending of their run streak compared to those with a less firm attachment. Greater individual attachment to a streak increased willingness of a runners to take measures to maintain their streak while enduring pain.

> *“I couldn’t go more than a mile or two and it would be like 10 or 11 minutes a mile because I was just in such pain… Just to sleep at night I had to take a ton of ibuprofen. I was in bed sleeping, it hurt. So I finally just you know, I finally after probably at least a month, maybe two months of this. That’s how, a bit obsessive I was, but I finally went to the doctor.”* Participant 10, Male.

When engaging in cessation of a run streak, the degree of attachment to the streak made this process more or less difficult. When streaking was a well-established component of a person’s identity and a part of their daily lives, ‘letting go’ of the streak became increasingly challenging.

> *“I guess it becomes a part of you, like this is who I am. This is what I’ve done. I don’t want to give it up now. It turned into I’m going to do it until I physically can’t or, you know, there’s a good reason not to.”* Participant 9, Female.

Consequently, when terminating a streak that a runner experienced an attachment to, the consequences were more severe than if attachment was low.

> *“When I was on my first big one, I would post about it, I felt like maybe it became more of my personality than it should have been. And then in turn, when that one had to break, it was much harder for me because I made such a big deal out of it. So now, I’m on my third of a decent length and I’ve just, you know, it is what it is. I do it if I can.”* Participant 14, Female.

### The Aftermath

During the period following cessation of long-term streaks runners reported feeling an immediate impact and most runners remembered and recalled this time clearly and in detail. The two sub-themes identified were: ‘*emotional cocktail*’, and ‘*reflexive rebound intentions*’.

#### Emotional cocktail

Immediately after breaking a streak, many runners reported experiencing a multitude of emotions of higher and lower activation including ‘sadness’, ‘frustration’, and ‘anger’, often simultaneously or sequentially. Runners experiencing abrupt and unexpected endings to their streak, due to reasons such as sickness or injury, reported the emergence of emotions almost immediately after ending their streak. Often the ending of the streak was experienced within an already adverse context such as during periods of pain, injury or health problems, leading to runners experiencing a ‘double downer’ of a broken streak and having to deal with other additional issues. When runners were forced to end their streak because of an upcoming surgery or medical event, they often dealt with emotions such as sadness upon receiving the news of the inevitable ending of their streak.

> *“I think it was harder the day I found out I couldn’t run. Like when I had the consultation with the surgeon, I remember calling my husband after I had the appointment and just like crying on the phone because I was really upset [about losing the streak]”.* Participant 17, Female.

Runners also reported emotions such as anger when recalling the end of their run streak. Anger was often directed at the individual themselves.

> *“Oh my gosh, I was so mad at myself for letting that happen.”* Participant 13, Female.

The most common emotion associated with ending a streak that was reported by individuals was disappointment.

> *“I felt disappointed immediately, right away.”* Participant 3, Female.

Intensity of these feelings were related to streak attachment. The greater the attachment to the streak, the more intense the emotional response seemed to be. In some instances, the ending of a streak provoked an emotional response wherein the runner cried.

> *“So I just called my wife, a bit tearfully honestly, I said, you know, I just, I can’t do it anymore, I have to realize it’s over.”* Participant 10, Male.

Consequently, runners who were less attached to their streak experienced less intense emotions than those with greater attachment.

> *“I did not cry myself to sleep or anything like that. It was mildly disappointing, but it wasn’t a huge, big, catastrophic disaster to me.”* Participant 5, Female.

Not all participants reported feelings of distress or recounted emotional states upon the breaking of their long-term streak. Some runners reported feeling at ease when their run streak ended.

> *“I put a lot of pressure on myself during the first streak and I was relieved that it was over.”* Participant 17, Female.

When runners felt burdened by their streak and experienced ‘streak stutters’ for a period of time, streak cessation could bring feelings of relief.

> *“It was also a relief. I was glad to not have to get up every day and run.”* Participant 9, Female.

Feelings of relief often did not exist in isolation. When runners reported feeling relieved upon streak cessation, it was often reported in combination with contrasting emotions.

> *“It was a little bit of a relief and a little bit of a let down at the same time. You’re just kind of like, you know, finally I can give my body a break. But at the same time, it’s like, you’ve gone 366 days like and then in one day you can just start the clock back from one if you decide to do it again.”* Participant 6, Other Gender.

#### Reflexive rebound intentions

Following streak cessation, runners reported various stances on their intents for future streaking. Most runners reported that directly following streak cessation, they intended to start another streak as soon as physically possible.

> *“But I also knew in my mind that I’d streak again. You know, that I’d get back up on that horse that threw me off.”* Participant 1, Male.

An immediate intention to streak again was reported to serve as a method of coping with the end of a streak. When runners were confident in their ability to restart a streak after it ended, there seemed less emotion associated with its ending. A future-oriented perspective seemed to help runners deal with the ending of their streak, particularly for those who had previous experiences of ending long-term streaks in the past.

> *“There’s a little sadness, but I’d already done it once before. I just knew for me, it’s kind of going to have to be my lifestyle. It just works for me. So if it stops, then when it’s ready to restart then we’ll just go again.”* Participant 14, Female.

For some, the initial intention to restart their streak was present after breaking their run streak but did not translate into action subsequently. The intention-behaviour gap was often justified by the enjoyment of ‘rest days’.

> *“I sort of thought I might start another longer one and then just never did […] I decided I like the days off.”* Participant 12, Male.

### The Adaptations

In the weeks and months after streak cessation, runners reported various personal impacts. Within the adaptations, three subthemes were identified: ‘*grief’, ‘return to running’* and *‘life lessons learnt’*.

#### Grief

Many runners reported going through a series of emotional and behavioural responses after the loss of their run streak. Some runners described what they referred to as a ‘grieving process’ where feelings of sadness and regret were experienced for a period of time.

> *“There was a sense of maybe I should have just ran. I should have just ran that day. I could have done it. I should have done it.”* Participant 2, Male.

During the grieving process, runners reported receiving social emotional support from friends and family during the days and weeks following the end of their streak.

> *[When recalling the days and weeks after termination of their streak] “So I had a ton of people just reaching out left and right.”* Participant 9, Female.

Over time, runners reported that the intensity of these feelings decreased, and they were able to become more accepting of the end of their streak.

> *“It’s intense at first and it, it is kind of like a grieving process where you become more accepting of it over time.”* Participant 4, Male.

However, some runners reported the re-emergence of sadness and regret near what would have been a ‘streaking milestone’.

> *“Sometimes I get a little sad that it’s over… I think about when we get to December 31^st^, when it would have been my 11 year anniversary, I think that will be kind of hard for me just because I’m like man, I could have kept on.”* Participant 11, Male.

Re-emergence of feelings of grief were also reported to occur when the runner was subjected to an environment in which they used to run streak. This was typically when runners seemed to have a strong streak attachment.

> *“It [feelings post-streak] hasn’t completely gone away, especially like a beautiful fall day, […], sunny, no wind, and I feel like geez I should just go out for a run.”* Participant 10, Male.

Not all runners reported a grieving process, particularly those who were less attached to their streak, and when finding other non-running related goal to pursue during the time one would typically engage in running.

> *“I adapted very quickly. After the injury […], I didn’t run a step for 6 weeks. […] I didn’t particularly miss it. I found other things to do.”* Participant 5, Female.

All three runners who voluntarily made the decision to end their streak in the absence of injury did not report experiencing a grieving process.

> *“I expected regret and got none […] It was fun. It was good. It was a fine number, but I moved on.”* Participant 12, Male.

#### Return to Running

Following cessation of a run streak, all runners who were physically capable eventually undertook regular running as an ongoing behaviour. The time between the end of a streak and the initiation of running varied. For some, this length of time was reported to be short.

> *“I think I was interrupted for two days. It might have been three, but it was no more than three. Right back into it.”* Participant 16, Male.

Other runners reported larger gaps in running behaviour following streak cessation.

> *“It was about a year and two months that I went between the first week and then starting the second one. And I don’t think I really ran once in that time.”* Participant 17, Female.

Although all physically capable runners began running regularly, seven opted not to initiate another run streak. Reasons for not re-engaging with run streaking included pursuing different goals, including running and non-running related ones.

> *“No, I would consider it [starting another run streak], I think mostly I just have other life plans and other interests.”*, Participant 7, male.

Runners who did not begin another streak also reported benefits of regular rests from an injury perspective and the enjoyment of having increased flexibility with their time and schedule.

> *“I’m enjoying the freedom of not being burdened of having to do it every day.”* Participant 15, Male.

However, most runners reported the initiation of a subsequent run streak following cessation of the previous one.

> *“I did know that I liked doing that and as soon as I was able to, I started another streak and got to where I am now.”* Participant 4, Male.

The decision to streak again was a personalized choice. However, an individual’s ‘love for running’ and their sense of identity associated with run streaking were common reasons for the decision to start another run streak.

> *“Running had become a very important part of who I am. It became like an identity thing. And being a streak runner became an identity thing, so I knew at some point I was gonna get back into it.”* Participant 2, Male.

In beginning another run streak, it was common for runners to reinitiate running by engaging in the behaviour a few times a week before starting another streak.

> *“So three months later is when I started running again. So I didn’t necessarily start streaking again. I think I’d been running two or three or four weeks before I started a streak again.”* Participant 4, Male.

#### Life lessons learnt

Several runners reported that run streaking made them better equipped to deal with adversity in other areas of their lives. Common lessons reported by streak runners who had ended a streak were characterized by concepts such as ‘grit’ and ‘mental strength’.

> *“I learnt a lot about myself and like pushing my limits as far as like, the brain is stronger than the body.”* Participant 11, Male.

Some runners reported learning about their own personal limits, concluding that they perceived themselves as more capable than they had previously thought.

> *“You learn a lot about mental strength. Because doing a run streak, training for races, you’re pushing yourself beyond what you’ve done before. When you get to the other side of that, you really understand that your boundaries aren’t always where you thought they were.”* Participant 6, Other Gender.

For some, run streaking was a source of self-efficacy. Runners found confidence in their own abilities through run streaking.

> *[When asked about lessons learnt from run streaking] “You can do a lot more than you ever thought you could. If you put your mind to it, you can do things that you never imagined you could do.”* Participant 16, Male.

## DISCUSSION

### Principal findings

Streak runners who ended a long-term run streak reported experiencing some unintended negative consequences of run streak cessation. Prior to streak cessation, many runners continued running for long periods despite injury. Streak runners were often willing to endure adverse consequences (e.g., pain) for long periods of time to maintain their run streak. Immediately following run streak cessation, runners reported a variety of affective responses including sadness, anger, disappointment, or relief. The intensity of responses seemed related to the level of streak attachment. Following streak cessation, many runners reported an immediate intention to streak again, often to cope with and to compensate for the loss of a previous streak. Ongoing and persistent feelings of grief following streak cessation were reported by some runners with stronger streak attachment and streaker identity. Despite some perceived negative consequences prior to and after streak cessation, no long-term negative consequences of streaking were observed in this study. Positive long-term effects included the maintenance of regular running behaviour following streak cessation, regardless of whether runners began streaking again. Most physically capable runners re-engaged in run streaking, with others engaged in running approximately 3-4 days per week. Most runners reported that run streaking taught them lessons about mental strength, grit, and pushing their limits. Several runners indicated that run streaking made them more capable to deal with adversity in various aspects of their lives. No runners reported regretting having engaged in long-term run streaking and all runners reported an overall net positive experience of streak running.

### Strengths and weaknesses of the study

Few studies examine the backfire potential of engaging in BCTs (27). This study provides an in-depth examination of the potential negative consequences of engaging in long-term run streaking in individuals whose long-term streak ended, with most runners reporting involuntary streak cessation. This study includes a population that demonstrates long-term engagement in a health behaviour using a specific BCT, which is rare. Focusing on runners who ended a long-term run streak of ≥100 days ensures that this study examines a population that is most likely to have experienced unintended negative consequences of the use of run streaking. When interpreting the findings it should be noted that most runners were from North American, Caucasian, and only one participant fell into the 20–29-year-old age group. Moreover, accounts of streak cessation were provided in retrospect, and the accuracy of participant accounts may be skewed over time.

### Interpretation of findings and relation to other studies

Long-term streaking may lead runners to develop a relationship with their streak as if it is a (living) thing. Consequently, the streak needs to be ‘kept alive’ and requires care and attention so that the streak does not die. Ongoing investment of energy into streak maintenance can lead to runners developing an attachment to their streak over time. The need to invest time and effort into the streak, and its ability to grow and age likely contributes to its biomorphized and (sometimes anthropomorphized) conceptualisation. In this state, the cessation of the streak constitutes not just an absence of behaviour, but a loss of a relationship with a (living) thing.

Initial evidence of the backfiring potential of using streaking as a BCT were highlighted in a study examining the experience of run streaking in recreational runners (26). The findings suggested that streaking can lead to lack of recovery, stress-inducement, and negative affective states when streaks break (26). The current study on runners ended a long-term run streak support these findings. Potential unintended negative consequences reported by some runners in the current study included a willingness to run with injury and endure pain while engaging in streaking. In the context of the avoidance-endurance model of pain (32), streak runners can be viewed as engaging in task persistence behaviours (e.g., running through injury) which carry the risk of pain chronification (33). As another unintended consequence, streak runners reported experiences such as sadness, anger, disappointment, regret and grief following streak cessation. These reports were not uniform and differed across streak runners. No evidence of long-term negative psychological consequences associated with run streaking were found.

Excessive engagement in running has raised questions of a potential upper threshold for health and longevity benefits of this behaviour (7). Some studies suggest that excessive running, may cause adverse effects on cardiac structure and function (34). However, results from a longitudinal study (n=55,137) comparing weekly running time and death rates showed no increased risk of all-cause mortality even in the highest running groups and suggests that insufficient evidence exists to conclude whether large amounts of running have adverse health consequences.(7).

In the self-enactable compendium (21), BCTs were supplemented with their associated potential **‘**possible adverse effects’ based on expert review of each of the techniques. Adverse effects such as ‘possible addiction’ (#62 pharmacological support) or feelings of unpleasantness and stress (#91 self-disincentive) were flagged when BCTs were used an inappropriate manner. The compendium highlights the need for a deeper analysis of BCT backfire potential and strategies to combat the negative consequences of BCT use. The current study provides information on the drawback effects and backfire potential of run streaking as a BCT.

The importance of learning from behaviour change interventions which have unintended negative consequences is often overlooked (35). Osman et al. (2020) proposed a taxonomy of failures of behavioural change examining conditions under which behaviour change techniques might lead to backfiring effects. Out of the eight potential types of failures listed in the taxonomy, the current research aligns with ‘treatment offset by negative side effects’. Within this category, the target behaviour of an intervention is achieved, however it coexists with negative consequences. In the context of ‘naturally occurring’ run streaking, the target behaviour (daily running) may be attained, however the unintended negative consequences (e.g., injury, risk of pain chronification) have the potential to outweigh the positive effects, demonstrated by for example runners’ reports of streak inconveniences. When the negative consequences become too great behavioural disengagement followed. However, the current study demonstrate that the unintended negative consequences of run streaking were often not sufficient for the behaviour to be abandoned voluntarily for prolonged periods of time. Moreover, the most prominent unintended negative consequences were often reported following run streak cessation. Side effects of discontinued BCT use included sadness, anger, disappointment, and grief. When individuals were no longer able to engage in the target behaviour of running every day, they experienced negative psychological consequences. However, regardless of negative consequences during and after engaging in run streaking, runners reported an overall net positive experience of streak running.

### Implications and future research

Few instances of voluntary cessation were examined in the current study, where three out of 17 runners voluntarily ended their run streak. Thus, whether there is a difference in the impacts of unintentional compared to voluntary streak cessation could not be examined. The three runners engaging in voluntary streak cessation reported minimal unintended negative consequences. However, further research on voluntary streak cessation is required.

This study found that runners with a high level of streak attachment had high levels of psychosocial unintended negative consequences following streak cessation. Future research might focus on examining individual differences in the level of streak attachment and the underlying motives for engaging in streak running as potential explanatory factors for differential perceived consequences of streak run cessation.

Additional questions of who might benefit from streaking remain. This study demonstrated that unintended negative consequences when run streaking were not uniform across participants. Future research should examine whether the technique is more suitable for certain individuals with different personality characteristics (e.g., openness, conscientiousness) or living contexts. Streaking as a BCT has the potential to influence other behaviour related cognitions and behaviours. A previous study noted ‘streaking spillover’ of using the technique to other areas of life (26). The current study confirmed these findings in streak runners who ended a long-term streak. Future research would benefit from systematically examining streaking drawing on evidence and theory focused on multiple behaviour change (36).

### Conclusion

Run streaking as a BCT has the potential to support long-term behaviour change like few other individual techniques. Runners become attached to their ever-growing run streak and consistently engaging in the behaviour to ‘keep the streak alive’. However, the aspects that contribute to streaking’s ability to support change also have the potential to cause adverse consequences such as inconvenience and a willingness to engage in pain endurance behaviours such as running through injury. Streak cessation can be associated with negative emotions which have the potential to develop into experiences of grief. The degree to which these factors impacted runners was variable and often associated with their level of streak attachment.

Although, there were potential backfiring effects, streaking was generally a net positive experience for participants and seen as helpful in the maintenance of running behaviour. No long-term negative consequences were observed. Run streaking can be seen as a beneficial behaviour change technique to help individuals increase their health-related behaviours, when accounting for possible backfire effects.

## Data Availability

Data cannot be shared publicly because of the qualitative nature of the study. Data are available from the corresponding author upon reasonable request.

## DATA AVAILABILITY STATEMENT

The data that support the findings of this study are available from the corresponding author upon reasonable request.

## ACKNOWLEDGEMENTS

The authors would like to thank all participants in the study.

## INSTITUTIONAL REVIEW BOARD STATEMENT

The study was conducted in accordance with the Declaration of Helsinki and was approved by an Institutional Review Board/Ethics committee. See details under Methods.

## Appendix A Topic Guide

### Part 1: Running and streaking in general

Let’s start with some questions about your running in general and your streak running.

**1. For how long have you been running overall, not just run streaking?**
**2. Besides running, do you do any other types of physical activity at the moment?**

a. Have you always been running as an activity, or have you done other types of activity in the past?
**3. For run streaking specifically, how long have you been doing it?**

a. Are you currently still streaking?
b. [If still streaking] What was your longest ever streak? Is that your current streak?
**4. How did you get into run streaking?**

a. Was it planned or accidental?
**5. Do you think run streaking is a useful strategy for everyone who want to be physically active?**

a. If so, why and for whom?
**6. What are some of the positives of run streaking?**

a. Routine
b. Accomplishment
c. Community
d. Fitness
**7. What might be some of the negatives of run streaking?**

a. Injury
b. Impact on social life
c. Stress
d. Time and energy
e. Costs of equipment
**8. How did you cope with some of the negative consequences of streaking?**
**9. Were there times where you felt like you were obsessed with your streak?**
**10. Did maintaining your streak ever feel like a burden?**

### Part 2: Streak breaking or retiring the streak

In this second part I would like to ask you a few questions about the time when you stopped your run streak. Here I am interested in the last streak that stopped which was longer than 100 days?

**11. Did you stop your streak intentionally or was it not intentional?**

a. *If intentional*: What led you to this decision? Was this a sudden decision, or was it a process?
b. *If non-intentional*: What was the reason why had to stop?

i. Injury or surgery, Life event, Job or family commitment
**12. Can you remember the actual day that your streak stopped?**

a. [If yes] Could you tell me about what happened that day?
**13. How did you feel after your streak stopped?**

a. The next day, weeks, month
**14. How did your life change after stopping your streak?**

a. What was your relationship with physical activity like?
b. What did you do with your free time?
c. How did it affect your social life?
**15. Did you continue to run after your streak ended?**

a. [If yes] When did you start again? How frequently did you run?
b. [If no] Did you do any other type of physical activity instead?
**16. Did you start another streak? Why/why not?**

a. [If yes] When did you decide that you will start another streak? How much time was there between the end of streak and the beginning of your next streak?
**17. What lessons have you learnt from run streaking?**

a. About yourself, physical activity, accomplishing goals
**18. What else you would like to tell us about streaking that we haven’t asked about?**

Thank you for answering all my questions. We really appreciate you participating in this research study. Finally, I would like to ask you some brief questions about yourself.

1. **19. Demographic Questions.**

a. How old are you?
b. How would you describe your gender?
c. What is your relationship status?
d. Height/ Weight?
e. How would you describe your ethnicity?
f. What is your employment status?

